# Performance of Rapid Antigen Tests Based on Symptom Onset and Close Contact Exposure: A secondary analysis from the Test Us At Home prospective cohort study

**DOI:** 10.1101/2023.02.21.23286239

**Authors:** Carly Herbert, Biqi Wang, Honghuang Lin, Nathaniel Hafer, Caitlin Pretz, Pamela Stamegna, Seanan Tarrant, Paul Hartin, Julia Ferranto, Stephanie Behar, Colton Wright, Taylor Orwig, Thejas Suvarna, Emma Harman, Summer Schrader, Chris Nowak, Vik Kheterpal, Elizabeth Orvek, Steven Wong, Adrian Zai, Bruce Barton, Ben Gerber, Stephenie C Lemon, Andreas Filippaios, Kylie D’Amore, Laura Gibson, Sharone Greene, Sakeina Howard-Wilson, Andres Colubri, Chad Achenbach, Robert Murphy, William Heetderks, Yukari C Manabe, Laurel O’Connor, Nisha Fahey, Katherine Luzuriaga, John Broach, David D McManus, Apurv Soni

## Abstract

**Background:** The performance of rapid antigen tests for SARS-CoV-2 (Ag-RDT) in temporal relation to symptom onset or exposure is unknown, as is the impact of vaccination on this relationship.

**Objective:** To evaluate the performance of Ag-RDT compared with RT-PCR based on day after symptom onset or exposure in order to decide on ‘when to test’.

**Design, Setting, and Participants:** The Test Us at Home study was a longitudinal cohort study that enrolled participants over 2 years old across the United States between October 18, 2021 and February 4, 2022. All participants were asked to conduct Ag-RDT and RT-PCR testing every 48 hours over a 15-day period. Participants with one or more symptoms during the study period were included in the Day Post Symptom Onset (DPSO) analyses, while those who reported a COVID-19 exposure were included in the Day Post Exposure (DPE) analysis.

**Exposure:** Participants were asked to self-report any symptoms or known exposures to SARS-CoV-2 every 48-hours, immediately prior to conducting Ag-RDT and RT-PCR testing. The first day a participant reported one or more symptoms was termed DPSO 0, and the day of exposure was DPE 0. Vaccination status was self-reported.

**Main Outcome and Measures:** Results of Ag-RDT were self-reported (positive, negative, or invalid) and RT-PCR results were analyzed by a central laboratory. Percent positivity of SARS-CoV-2 and sensitivity of Ag-RDT and RT-PCR by DPSO and DPE were stratified by vaccination status and calculated with 95% confidence intervals.

**Results:** A total of 7,361 participants enrolled in the study. Among them, 2,086 (28.3%) and 546 (7.4%) participants were eligible for the DPSO and DPE analyses, respectively. Unvaccinated participants were nearly twice as likely to test positive for SARS-CoV-2 than vaccinated participants in event of symptoms (PCR+: 27.6% vs 10.1%) or exposure (PCR+: 43.8% vs. 22.2%). The highest proportion of vaccinated and unvaccinated individuals tested positive on DPSO 2 and DPE 5-8. Performance of RT-PCR and Ag-RDT did not differ by vaccination status. Ag-RDT detected 78.0% (95% Confidence Interval: 72.56-82.61) of PCR-confirmed infections by DPSO 4. For exposed participants, Ag-RDT detected 84.9% (95% CI: 75.0-91.4) of PCR-confirmed infections by day five post-exposure (DPE 5).

**Conclusions and Relevance:** Performance of Ag-RDT and RT-PCR was highest on DPSO 0-2 and DPE 5 and did not differ by vaccination status. These data suggests that serial testing remains integral to enhancing the performance of Ag-RDT.

## Introduction

Rapid antigen tests (Ag-RDTs) are commonly used to diagnose COVID-19 due to their availability over-the-counter for home use, relatively low cost, and ability to return results in 15-20 minutes.^1–3^ Previous work has informed the FDA guidance on *testing frequency* to minimize the risk of false negative tests in symptomatic as well as asymptomatic individuals.^4^ However, important questions remain about *when to begin testing*, particularly among those with symptoms or after close contact with an infected person.^4,5^ Similarly, our understanding of Ag-RDT performance by time past exposure is limited.^6–8^ Understanding Ag-RDT performance change over the symptom course and in relation to SARS-CoV-2 exposure is crucial to guide optimal use of diagnostics for risk assessment.

Several prior studies have examined Ag0RDT performance when used serially, but these studies predate the arrival of the Omicron variants in the United States and widespread vaccination coverage.^9,10^ Currently, approximately four in every five U.S. adults have received at least one dose of a SARS-CoV-2 vaccine.^11^ Vaccination has been associated with changes in the signs and symptoms of SARS-CoV-2 infection, including fewer symptoms and a higher likelihood of asymptomatic infections.^12^ The performance of molecular diagnostics, including reverse transcriptase polymerase chain reaction (RT-PCR), and Ag-RDTs is closely related to detectable viral load; therefore, it is important to determine whether changes in viral dynamics and symptomology due to vaccination have an impact on diagnostic performance.

Using data from Test Us at Home, a prospective cohort study that enrolled participants from throughout the United States, we examined paired serial Ag-RDT and molecular testing to determine how relative sensitivities of Ag-RDT and RT-PCR tests vary by day past symptom onset and exposure and how these findings vary based on vaccination status. The results of this study will inform pragmatic use of Ag-RDT at-home tests to detect SARS-CoV-2.

## Methods

### Study Population

In this analysis, we used data from the Test Us at Home study, a longitudinal cohort study that evaluated the performance of serial use of Ag-RDTs for detection of COVID-19 among asymptomatic individuals.^4^ The Test Us at Home study enrolled participants ages 2 years and older across the United States between October 18, 2021 and February 4, 2022. This study was approved by the WIRB-Copernicus Group (WCG) Institutional Review Board (20214875). Only participants who completed at least one Ag-RDT or RT-PCR were included in this analysis. Participants were included in the Day Post Symptom Onset (DPSO) analyses if they self-reported any symptoms during the study period (Figure 1). Participants who had a RT-PCR+ result more than 14 days before or after symptom onset were excluded, as these symptoms were assumed to be unrelated to the observed infection.^13^ Participants who reported experiencing a COVID-19 exposure during the study were included in the Day Post Exposure (DPE) analysis. Participants with an index RT-PCR+ result more than 14 days after the reported exposure were excluded from the DPE analyses.

**Figure 1:**
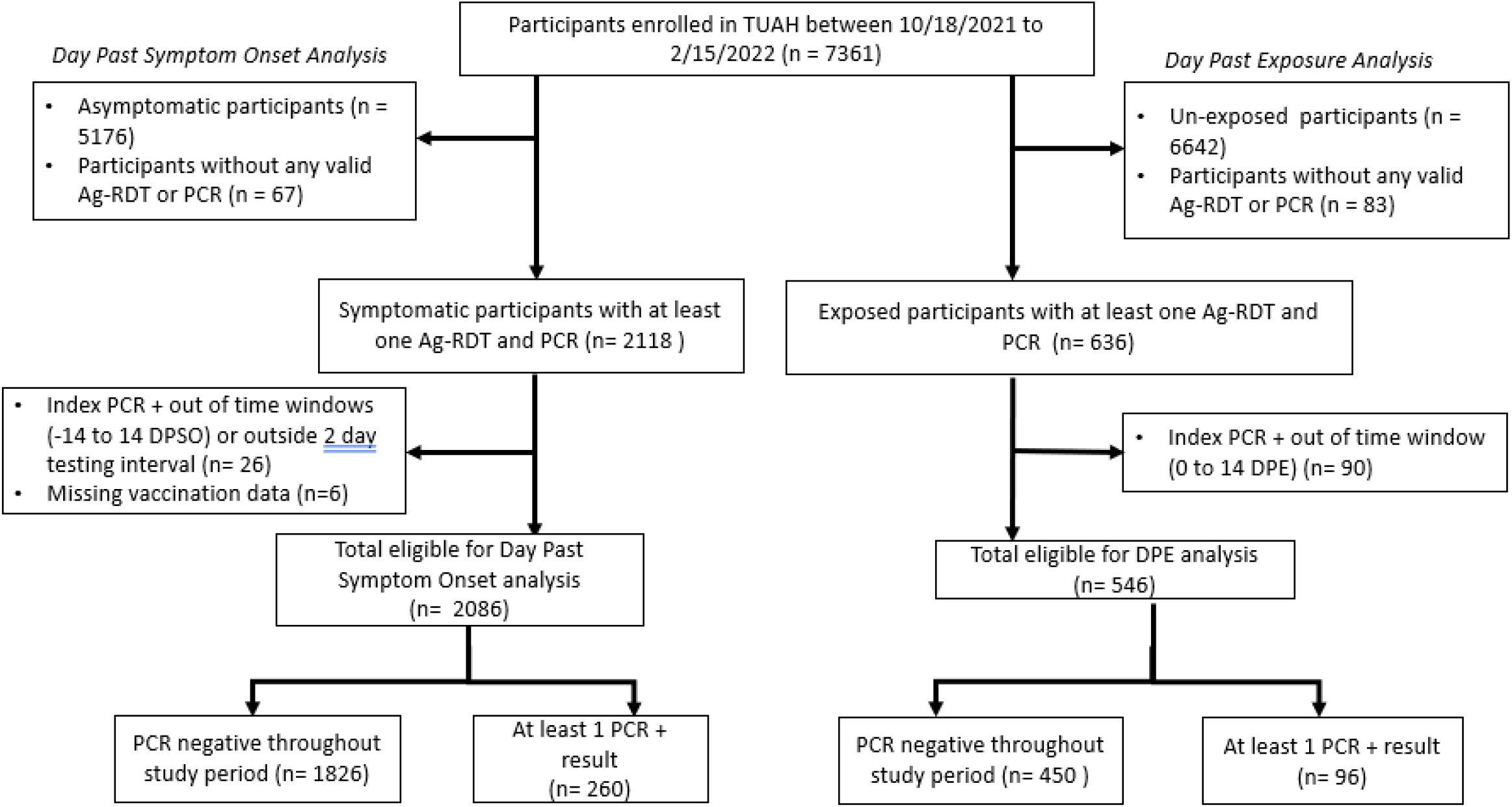
Consort Diagram for DPSO and DPE Analyses.

All Test Us at Home participants were asked to conduct Ag-RDT and RT-PCR testing every 2 days over a 15-day period. Participants were asked if they had any symptoms in the last 48 hours or any known exposures to SARS-CoV-2 at the beginning of each testing session. During each testing session, two anterior nasal swabs were collected by the participant; one swab was used for performing an Ag-RDT at home, while the other swab was sent to a central laboratory for RT-PCR testing. All study activities and questionnaires were conducted through a custom study app. Additional details about the study design, protocol, and participants are described elsewhere.^4,14^

### Measures

#### Symptoms

Participants self-reported symptoms (fever, body aches, fatigue, rash, nausea, abdominal pain, diarrhea, loss of smell, runny nose, cough, headache, or other) during each testing period immediately prior to using the Ag-RDT (every 48 hours). The first day that a participant reported 1 or more symptoms was termed DPSO 0.

#### Close-Contact Exposure

Participants self-reported close-contact exposures to COVID-19 at the time of baseline study enrollment and before each testing period. An exposure was defined as being within 6 feet of an infected person without a mask for at least 15 minutes over a 24-hour period. DPE 0 was defined as the first day of the reported exposure.

#### Vaccination Status and Previous Infections

Vaccination status and history of previous SARS-CoV-2 infection was self-reported during the enrollment survey. Vaccination status was operationalized into two groups: vaccinated (V1 dose) and unvaccinated (0 doses). Previous infections with SARS-CoV-2 were self-reported.

#### Molecular Testing (RT-PCR)

Molecular comparator RT-PCR results were based on a combination of molecular test results for the detection of SARS-Cov-2 infection.^4^ Cycle threshold (Ct) values for the E-gene from RT-PCR were used to quantify viral load.

#### Rapid antigen test positivity

Participants were asked to provide an interpretation of each Ag-RDT (positive, negative, or invalid) and upload a picture of the test result to the study app. All self-reported positive tests were confirmed by study coordinators using the uploaded images.

#### Data Analysis

Percent positivity and cumulative positivity of all symptomatic and/or exposed participants were calculated for RT-PCR and Ag-RDT by DPSO and DPE and stratified by vaccination status with 95% confidence intervals using Wilson’s method.^15^ Cumulative positivity was defined as the number of participants with at least one positive test result over the number of participants who had taken at least one test until each DPSO and DPE. Sensitivity analysis was performed to assess whether findings differed between partially vaccinated (1 dose) and fully vaccinated (2+ doses) participants. All analyses were conducted using R software package version 4.2.1.

## Results

### Characteristics of Symptomatic and Exposed Participants

A total of 2,086 of the 7,361 total Test Us At Home participants (28.3%) reported symptoms associated with COVID-19 infection and were eligible for the DPSO analysis; 546 participants reported a close-contact exposure and were eligible for the DPE analysis (Figure 1). Approximately 10% of vaccinated and 20% of unvaccinated participants reported at least one previous SARS-CoV-2 infection (Table 1). Participants less than 18 years old comprised 36.9% and 47.6% of unvaccinated participants in the DPSO and DPE analyses, respectively, while they comprised 7.2% and 10.6% of vaccinated individuals. The majority of vaccinated participants (1677, 93.8%) had received 2+ doses of a SARS-CoV-2 vaccine.

**Table 1:** Characteristics of Vaccinated and Unvaccinated Participants included in Analyses

|  |  | Included in DPSO analysis<br>(n= 2086) |  | Included in DPE analysis<br>(n=546) |  |
| --- | --- | --- | --- | --- | --- |
|  |  | Vaccinated<br>(n=1788) | Unvaccinated<br>(n=298) | Vaccinated<br>(n=464) | Unvaccinated<br>(n=82) |
| **Confirmed RT-PCR+:**<br>**n (%)** |  | 179 (10.0) | 81 (27.2) | 64 (13.8) | 32 (39.0) |
| **Symptomatic: n (%)** |  | 1788 (100.0) | 298 (100.0) | 233 (50.2) | 45 (54.9) |
| **Previous Infections** |  |  |  |  |  |
|  | None | 1608 (89.9) | 235 (78.9) | 419 (90.3) | 64 (78.0) |
|  | 1 | 165 (9.2) | 57 (19.1) | 38 (8.2) | 17 (20.7) |
|  | 2+ | 15 (0.8) | 6 (2.0) | 7 (1.5) | 1 (1.2) |
| **Participant Age**<br>**(years): n (%)** |  |  |  |  |  |
|  | <18 years | 128 (7.2) | 110 (36.9) | 49 (10.6) | 39 (47.6) |
|  | 18-44 | 1158 (64.8) | 140 (47.0) | 272 (58.6) | 31 (37.8) |
|  | 45-64 | 401 (22.4) | 43 (14.4) | 129 (27.8) | 10 (12.2) |
|  | ≥65 years | 101 (5.6) | 5 (1.7) | 14 (3.0) | 2 (2.4) |
| **Gender: n (%)** |  |  |  |  |  |
|  | Men | 458 (25.6) | 93 (31.2) | 145 (31.2) | 34 (41.5) |
|  | Women | 1263 (70.6) | 199 (66.8) | 309 (66.6) | 45 (54.9) |
|  | Transgender | 4 (0.2) | 0 (0.0) | 0 (0.0) | 0 (0.0) |
|  | Non-binary | 51 (2.9) | 1 (0.3) | 9 (1.9) | 0 (0.0) |
|  | Missing | 12 (0.7) | 5 (1.7) | 1 (0.2) | 3 (3.7) |
| **Race: n (%)** |  |  |  |  |  |
|  | White | 1451 (81.2) | 252 (84.6) | 361 (77.8) | 72 (87.8) |
|  | Asian | 84 (4.7) | 5 (1.7) | 32 (6.9) | 1 (1.2) |
|  | Black/African-American | 49 (2.7) | 14 (4.7) | 14 (3.0) | 3 (3.7) |
|  | Native American/Alaskan Native | 4 (0.2) | 1 (0.3) | 3 (0.6) | 0 (0.0) |
|  | Native Hawaiian or Pacific Islander | 11 (0.6) | 1 (0.3) | 0 (0.0) | 0 (0.0) |
|  | Multiracial | 102 (5.7) | 8 (2.7) | 19 (4.1) | 1 (1.2) |
|  | Other | 63 (3.5) | 9 (3.0) | 27 (5.8) | 5 (6.1) |
|  | Missing | 24 (1.3) | 8 (2.7) | 8 (1.7) | 0 (0.0) |
| **Hispanic: n (%)** |  | 100 (5.6) | 33 (11.1) | 28 (6.0) | 10 (12.2) |
| **Education Level: n (%)** |  |  |  |  |  |
|  | Bachelor's Degree or higher | 1120 (62.6) | 50 (16.8) | 266 (57.3) | 14 (17.1) |
|  | Some college | 395 (22.1) | 79 (26.5) | 106 (22.8) | 16 (19.5) |
|  | High school graduate | 131 (7.3) | 57 (19.1) | 39 (8.4) | 13 (15.9) |
|  | Did not finish high school | 107 (6.0) | 78 (26.2) | 40 (8.6) | 32 (39.0) |
|  | Don't know | 12 (0.7) | 15 (5.0) | 4 (0.9) | 5 (6.1) |
|  | Missing | 23 (1.3) | 19 (6.4) | 9 (1.9) | 2 (2.4) |
| **Employment Status: n (%)** |  |  |  |  |  |
|  | Working now | 1208 (67.6) | 108 (36.2) | 342 (73.7) | 23 (28.0) |
|  | Student | 294 (16.4) | 115 (38.6) | 78 (16.8) | 43 (52.4) |
|  | Retired | 99 (5.5) | 3 (1.0) | 9 (1.9) | 2 (2.4) |
|  | Unemployed | 169 (9.5) | 63 (21.1) | 27 (5.8) | 14 (17.1) |
|  | Other | 7 (0.4) | 4 (1.3) | 2 (0.4) | 0 (0.0) |
|  | Missing | 11 (0.6) | 5 (1.7) | 6 (1.3) | 0 (0.0) |

### Percent Positivity of Ag-RDT and Molecular Tests by Day Past Symptom Onset among Vaccinated and Unvaccinated Individuals

Among 2,086 participants that reported at least one symptom, 12.5% tested positive by RT-PCR during the study (Table 1; Supplemental Figure 1a). Significantly fewer symptomatic vaccinated participants tested positive for SARS-CoV-2 during the study period compared to unvaccinated participants (Unvaccinated: PCR+: 27.6% vs Vaccinated PCR+: 10.1%; Unvaccinated Ag-RDT+: 25.3% vs. Vaccinated Ag-RDT+: 10.9%) (Supplemental Figure 1b). Trends were similar among those who were fully vaccinated (2+ doses) and partially vaccinated (1 dose) (Supplemental Figure 1c). The highest proportion of vaccinated and unvaccinated symptomatic individuals tested positive on DPSO 2 (Figure 2a, 2b).

**Figure 2:**
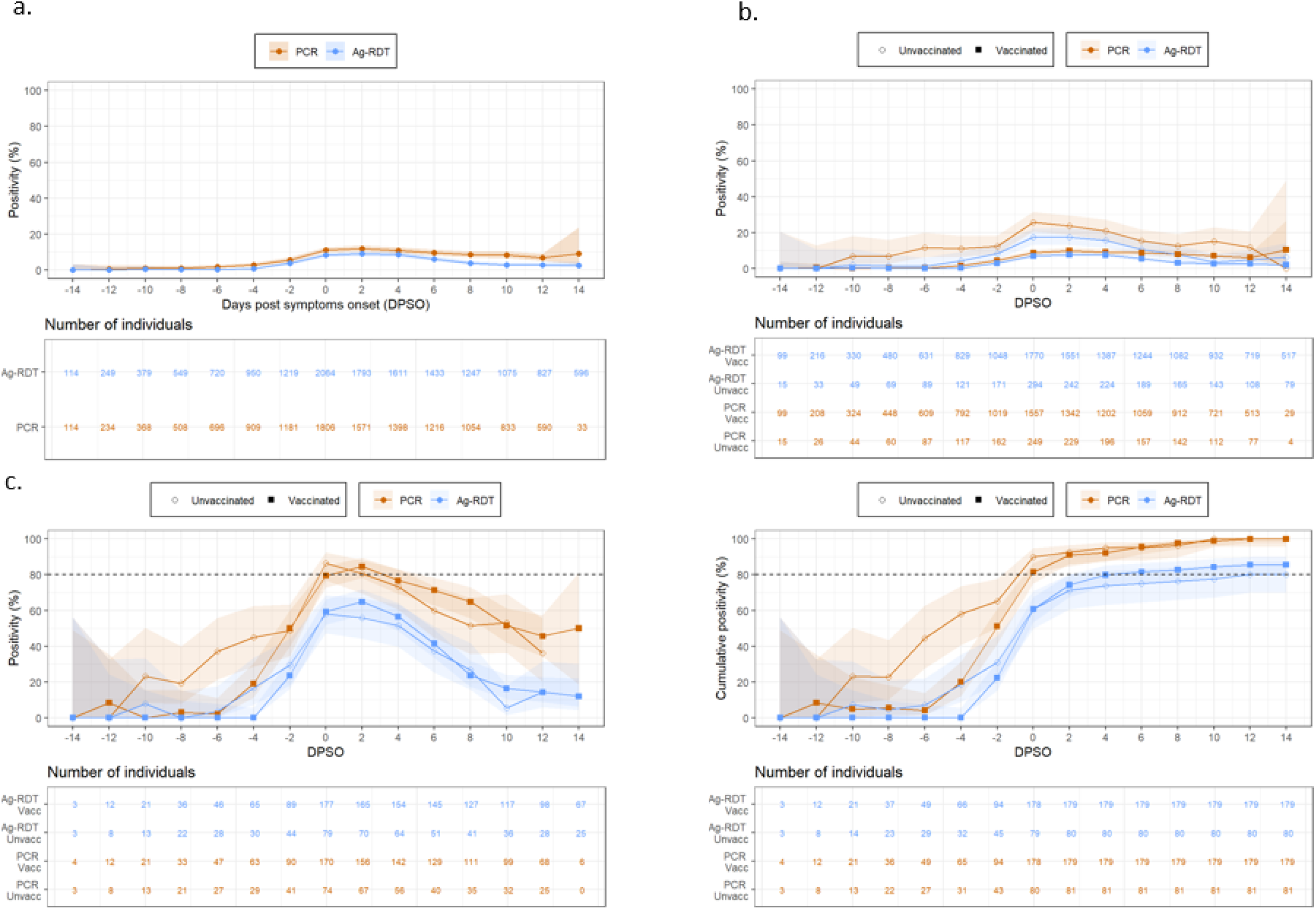
PCR and Rapid Antigen Test Positivity by Day Past Symptom Onset.

### Percent positivity of Rapid Antigen Tests and Molecular Tests by Day Past Exposure among Vaccinated and Unvaccinated Individuals

Of the 546 participants who reported a close-contact exposure during the study, 17.6% tested positive by RT-PCR (Table 1; Supplemental Figure 1d). More than 50% of vaccinated and unvaccinated individuals with close contact exposures also reported COVID-related symptoms during the study (Table 1). Exposed unvaccinated participants tested positive for SARS-CoV-2 twice as often than exposed vaccinated participants, on both RT-PCR and Ag-RDT (Unvaccinated PCR+: 43.8% vs. Vaccinated PCR+: 22.2%; Unvaccinated Ag-RDT+: 44.3% vs Vaccinated Ag-RDT+: 23.4%) (Supplemental Figure 1e). The cumulative positivity of exposed participants was conditioned on symptom status, such that exposed individuals with symptoms were much more likely to test positive within the first week since exposure (≤DPE 6) than participants who were exposed but did not have symptoms on the day of testing (Supplemental Figure 2). Percent positivity was highest on DPE 5 through DPE 8, irrespective of vaccination or symptom status (Figure 3a, 3b).

**Figure 3:**
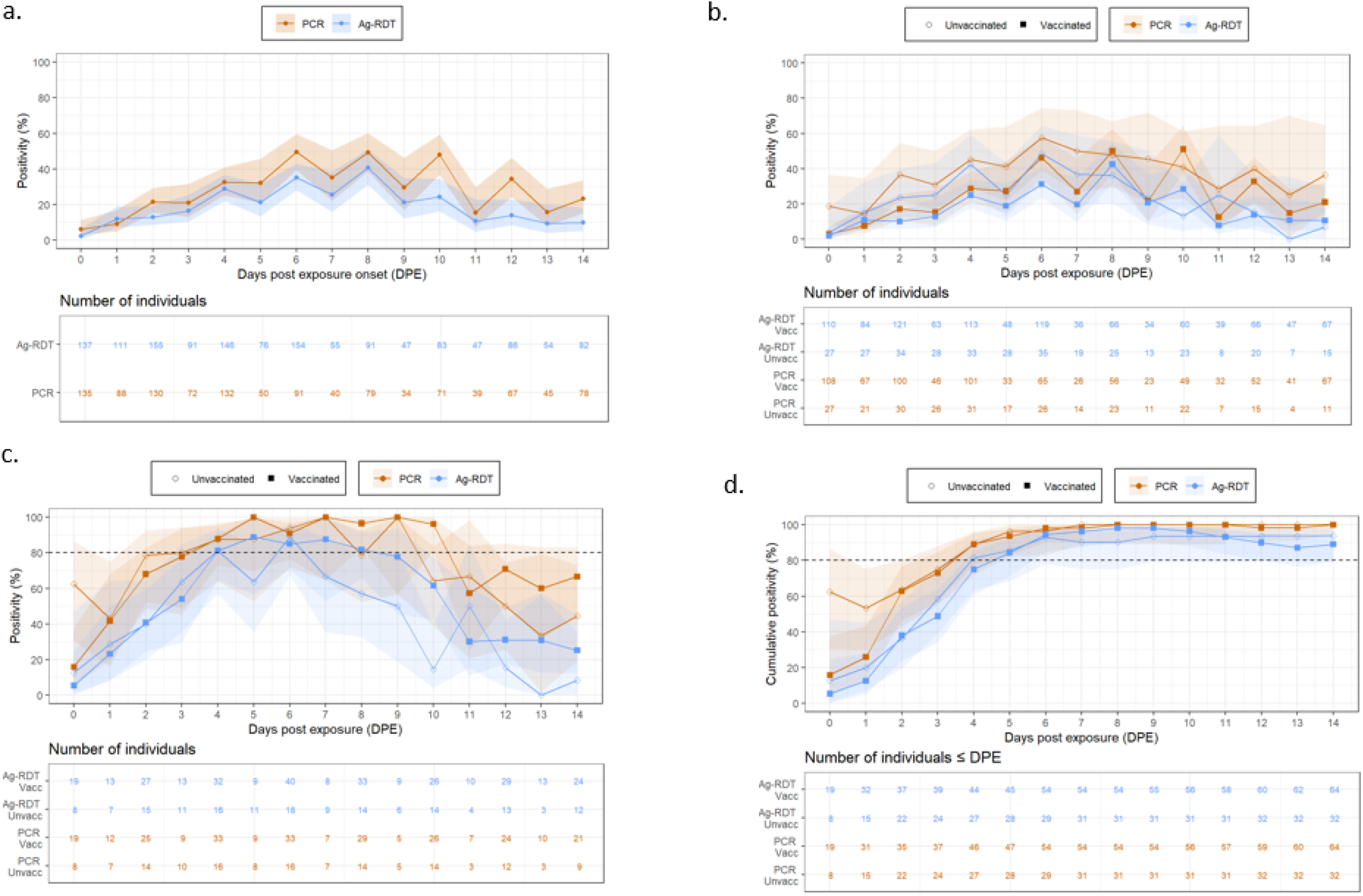
PCR and Rapid Antigen Test Positivity by Day Past Exposure.

### Diagnostic Performance by Timing of Symptom Onset and Exposure

We did not observe significant differences in the performance of RT-PCR and Ag-RDT by vaccination status by DPSO or DPE (Figure 2, Figure 3), and Ct values did not differ significantly between vaccinated and unvaccinated participants (Supplemental Figure 3). Among both vaccinated and unvaccinated participants, Ct value was lowest (i.e., highest viral burden) at DPSO 0-2 and DPE 5 (Supplemental Figure 3). Among symptomatic participants, RT-PCR detected 81.6% (95% CI: 76.2-85.9) of all PCR-confirmed infections on DPSO 0, while Ag-RDT detected 59.0% (95% CI: 52.9-64.8) of PCR-confirmed infections at this time (Figure 2c, 2d, Supplemental Table 1). Ag-RDT detected 79.9% (95% CI: 73.4-85.1) of vaccinated PCR-confirmed infections and 73.8% (95% CI: 63.2-82.1) of unvaccinated PCR-confirmed infections by DPSO 4 (Figure 2c). For exposed participants, RT-PCR and Ag-RDT detected over 94.7% (95% CI: 87.1-97.9) and 84.9% (95% CI: 75.0-91.4) of PCR-confirmed infections on day five post-exposure (DPE 5), respectively (Figure 3c, 3d, Supplemental Table 2).

## Discussion

In this study, we report the performance of nasal-swab Ag-RDT and molecular testing by time since symptom onset and time since exposure among individuals who were and were not vaccinated against SARS-CoV-2. Our study highlights three important findings: 1) we report that the real-world performance of Ag-RDT and RT-PCR peaked at DPSO 0-2 for symptomatic individuals and on DPE 5 for exposed individuals; 2) we show that the performance of Ag-RDT tests is similar among vaccinated and unvaccinated participants, likely because the viral peak did not differ between these groups; and 3) we demonstrate real-world evidence that participants who were vaccinated had significantly lower likelihood of testing positive on RT-PCR or Ag-RDT after an exposure than those who were unvaccinated. Taken together, these findings reinforce the importance of Ag-RDT tests for detection of SARS-CoV-2 virus and highlight the need for serial testing after symptom onset or exposure, as well as indicate the continued importance of vaccination.

As the pandemic enters its third year, use of COVID-19 diagnostics has shifted away from general screening and mandated testing towards personal risk assessment, with most people using Ag-RDT in response to acute symptoms or COVID-19 exposure.^16^ It is increasingly important to advise individuals on the timing of Ag-RDT use, to facilitate accurate test interpretation and minimize false-negative results. The present results reinforce the importance of serial testing when individuals are either symptomatic or exposed to SARS-CoV-2, in line with our previous recommendations (i.e., symptomatic individuals should perform two Ag-RDT 48 hours apart and asymptomatic individuals should perform three Ag-RDT 48 hours apart each).^4^ In case of symptoms, we recommend that individuals test with Ag-RDT on DPSO 2 and 4 to minimize the risk of false negative results. Further, to decrease transmission of SARS-CoV-2, individuals should isolate until DPSO 4, regardless of a negative test result on DPSO 2. For symptomatic individuals in our study, close to 80% of PCR-confirmed infections were detected by Ag-RDT by DPSO 4. Among exposed participants, Ag-RDT detected 5 in 6 PCR-confirmed infections by DPE 5, regardless of symptomatic status. Few new cases of SARS-CoV-2 were detected following DPE 6. These results were consistent among vaccinated and unvaccinated participants and support the CDC recommendations to isolate for 5 days post-exposure.^17^ A previous study of 225 individuals with PCR-confirmed SARS-CoV-2 infections similarly found that RT-PCR had a positivity rate of approximately 60% on the day of illness onset (defined as symptom onset among symptomatic individuals and first RT-PCR+ test among asymptomatic individuals).^10^ However, these investigators found that PCR-positivity and Ag-RDT-positivity peaked on day 3 and 4 past illness onset, which differs from our own results showing that that diagnostic performance peaked on DPSO 0 and 2. The observed difference may be explained by the emergence of new variants, as over 75% of infections in our study were due to the Omicron variant, which has a shorter incubation period than all previous SARS-CoV-2 strains.^18,19^ This discrepancy between results emphasizes the importance of re-evaluation of SARS-CoV-2 diagnostic performance and testing advisories as new SARS-CoV-2 variants continue to arise.

To our knowledge, most studies evaluating treatment effects of vaccination for SARS-CoV-2 has demonstrated vaccine’s efficacy for preventing severe disease, hospitalization, and death from COVID-19, but few have demonstrated efficacy for preventing infections. Previous studies have been limited in their ability to determine the impact of vaccination on susceptibility and transmission of COVID-19.^20–22^ In our study, unvaccinated individuals exposed to SARS-CoV-2 had nearly twice the likelihood of infection compared to exposed vaccinated individuals. This finding indicates that SARS-CoV-2 vaccination may indeed prevent against infection following exposure to the virus. This is especially noteworthy, as breakthrough infections due to the Omicron variant have become common.^23^ Our finding provides real-world data that is similar to previous reports based on passive data collection through electronic medical records and workplace studies, which suggest that even one dose of vaccination decreases the risk of infection from SARS-CoV-2. ^24–26^ In contrast to those studies, we account for both RT-PCR and Ag-RDT results in this study, and all participants, regardless of vaccination status, used the same diagnostics and procedures to screen for infection every 48 hours, which allowed for rigorous evaluation of the efficacy of vaccination for preventing infection after exposure. It is important to note that, among those who acquired infection, vaccination status did not affect RT-PCR or Ag-RDT tests’ sensitivity or viral dynamics, thus not requiring different testing strategies based on vaccination status.

Despite differences in the rates of infection among vaccinated and unvaccinated participants, once infected, peak viral load did not differ between vaccinated and unvaccinated individuals, consistent with previous reports. ^27,28^ This also helps to explain why we did not observe a significant difference in performance of Ag-RDT among vaccinated and unvaccinated individuals, as performance of Ag-RDTs is tightly correlated with viral load, with Ag-RDT diagnostic performance showing major declines when Ct > 30.^18^ Vaccinated and unvaccinated individuals did not differ in magnitude nor timing of viral load with relation to DPSO and DPE; therefore, no differences in diagnostic performance would be expected. However, while magnitude and timing of peak viral load did not differ between vaccinated and unvaccinated individuals, previous studies have shown that the duration of infectiousness may differ, with unvaccinated individuals showing prolonged infectiousness. Together, these results add to the evidence that vaccination against SARS-CoV-2 may decrease the risk of subsequent infection, but more studies are needed to understand the impact of vaccination on levels of infectiousness, as a function of viral load, during SARS-CoV-2 infection.

### Study Strengths and Limitations

This is one of the first studies to analyze the diagnostic performance of RT-PCR and Ag-RDT for COVID-19 based on days past acute symptom onset or exposure to an individual infected with SARS-CoV-2. Our study assessed serial paired longitudinal data to evaluate the performance of Ag-RDT and RT-PCR over the duration of infection using a large nationwide sample of children and adults. This is also, to the best of our knowledge, the first study to quantify time from exposure to Ag-RDT positivity.

Our study has limitations that need to be considered when interpreting our findings. Paired Ag-RDT and RT-PCR testing, as well as symptom trackers, were completed by participants every 48-hours. Assessing diagnostic performance at a finer temporal resolution might be useful in future studies. Symptoms, exposures, and Ag-RDT results were based on participant self-report. However, all positive Ag-RDT results were verified by research coordinators using images of the test strip that were uploaded by participants. In this analysis, we categorized anyone who received 1 or more vaccines for SARS-CoV-2 as vaccinated due to sample size limitations; therefore, there may be heterogeneity in the vaccine responses and immunity within this group. However, sensitivity analyses showed that results were consistent when those with 1 vaccination dose and 2+ doses were examined separately.

## Conclusions

In conclusion, this study supports testing immediately after symptom onset and between 3-5 days after exposure for optimal detection of SARS-CoV2 virus. Our findings suggest that vaccination prevents SARS-CoV-2 infection but not does not affect the performance of Ag-RDT or RT-PCR tests. Taken in sum, our results highlight the effectiveness of vaccination and serial testing in the context of symptom onset or close contact as public health strategies to manage transmission of SARS-CoV-2 virus.

## Data Availability

All data produced are available online at the NIH-RADx Data Hub.

https://radx-hub.nih.gov/

## Acknowledgment

We are grateful to our study participants and to our collaborators from the National Institute of Health (NIBIB and NHLBI) who provided scientific input into the design of this study and interpretation of our results, but could not formally join as co-authors due to institutional policies and to the Food and Drug Administration (Office of In Vitro Diagnostics and Radiological Health) for their involvement in the primary TUAH study. We received meaningful contributions from Drs. Bruce Tromberg, Jill Heemskerk, Felicia Qashu, Dennis Buxton, Erin Iturriaga, Jue Chen, Andrew Weitz, and Krishna Juluru. We are thankful to county health departments across the country who helped with recruitment for this siteless study by spreading the word in their networks.

