## Supplemental Tables for "Performance of Rapid Antigen Tests Based on Symptom Onset and Close Contact Exposure: A secondary analysis from the Test Us At Home prospective cohort study"

Supplemental Table 1: Performance of Ag-RDT and RT-PCR by Day Past Symptom Onset among RT-PCR Confirmed Participants

|  | Vaccinated Individuals | | Unvaccinated Individuals | | All Individuals | |
| --- | --- | --- | --- | --- | --- | --- |
| Day Past Symptom Onset (DPSO) | RT-PCR | Ag-RDT | RT-PCR | Ag-RDT | RT-PCR | Ag-RDT |
| -4 | 19.05  (11.25-30.41) | 0.00  (0.0-5.58) | 44.83  (28.41-62.45) | 16.67  (7.34-33.56) | 27.17  (19.14-37.04) | 5.26  (2.27-11.73) |
| -2 | 50.00  (39.88-60.12) | 23.60  (15.98-33.39) | 48.78  (34.25-63.52) | 29.55  (18.16-44.22) | 49.62  (41.19-58.07) | 25.56  (18.91-33.59) |
| 0 | 79.41  (72.72-84.81) | 59.32  (51.96-66.29) | 86.49  (76.88-92.49) | 58.23 (47.22-68.47) | 81.56  (76.22-85.92) | 58.98  (52.87-64.83) |
| 2 | 84.62  (78.13-89.44) | 64.85  (57.30-71.72) | 80.60  (69.58-88.29) | 55.71  (44.08-66.75) | 83.41  (77.97-87.72) | 62.13  (55.78-68.09) |
| 4 | 76.76  (69.16-82.95) | 56.49  (48.60-64.07) | 73.21  (60.41-83.04) | 51.56  (39.58-63.37) | 75.76  (69.33-81.20) | 55.05  (48.41-61.51) |
| 6 | 71.32  (62.99-78.42) | 41.38  (33.69-49.52) | 60.00  (44.60-73.65) | 37.25  (25.32-50.97) | 68.64  (61.30-75.15) | 40.31  (33.69-47.30) |
| 8 | 64.86  (55.62-73.11) | 23.62  (17.08-31.72) | 51.43  (35.57-67.01) | 26.83  (15.69-41.93) | 61.64  (53.55-69.14) | 24.40  (18.53-31.42) |
| 10 | 51.52  (41.80-61.12) | 16.24  (10.65-23.98) | 53.13  (36.45-69.13) | 5.56  (1.54-18.14) | 51.91  (43.42-60.29) | 13.73  (9.16-20.07) |
| Cumulative to DPSO 4 | 92.18  (87.30-95.28) | 79.89  (73.42-85.10) | 95.06  (87.98-98.06) | 73.75  (63.18-82.14) | 93.08  (89.32-95.58) | 77.99  (72.56-82.61) |
| Cumulative to DPSO 6 | 95.53  (91.43-97.72) | 81.56  (75.24-86.56) | 95.06  (87.98-98.06) | 75.00  (64.52-83.19) | 95.38  (92.11-97.34) | 79.54 (74.21-85.04) |
| Cumulative to DPSO 10 | 98.88  (96.02-99.69) | 84.36  (78.32-88.95) | 100.00  (95.47-100) | 77.50  (67.21-85.27) | 99.23  (97.24-99.79) | 82.24  (77.12-86.41) |

Supplemental Table 2: Performance of Ag-RDT and RT-PCR by Day Past Exposure among RT-PCR Confirmed Participants

|  | Vaccinated Individuals | | Unvaccinated Individuals | | All Individuals | |
| --- | --- | --- | --- | --- | --- | --- |
| Day Past Exposure (DPE) | RT-PCR | Ag-RDT | RT-PCR | Ag-RDT | RT-PCR | Ag-RDT |
| 0 | 15.79  (5.52-37.57) | 5.26  (0.27-24.64) | 62.50  (30.57-86.32) | 12.50  (0.64-47.09) | 29.63  (15.85-48.48) | 7.41  (2.06-23.37) |
| 2 | 68.00  (48.41-82.79) | 40.74  (24.51-59.27) | 78.57  (52.41-92.43) | 40.00  (19.82-64.25) | 71.79  (56.22-83.46) | 40.48  (27.04-55.51) |
| 4 | 87.88  (72.67-95.18) | 81.25  (64.69-91.11) | 87.50  (63.98-96.50) | 81.25  (56.99-93.41) | 87.76  (75.76-94.27) | 81.25  (68.06-89.81) |
| 6 | 90.91  (76.43-96.86) | 85.00  (70.93-92.94) | 93.75  (71.67-99.68) | 88.89  (67.20-96.90) | 91.84  (80.81-96.78) | 86.21  (75.07-92.84) |
| 8 | 96.55  (82.82-99.82) | 81.82  (65.61-91.39) | 78.57  (52.41-92.43) | 57.14  (32.59-78.62) | 90.70  (78.40-96.32) | 74.47  (60.49-84.75) |
| 10 | 96.15  (81.11-99.80) | 61.54  (42.53-77.57) | 64.29  (38.76-83.66) | 14.29  (4.01-39.94) | 85.00  (70.93-92.94) | 45.00  (30.71-60.17) |
| 12 | 70.83  (50.83-85.09) | 31.03  (17.28-49.23) | 50.00  (25.38-74.62) | 15.38  (4.33-42.23) | 63.89  (47.58-77.52) | 26.19  (15.30-41.07) |
| 14 | 66.67  (45.37-82.81) | 25.00  (12.00-44.90) | 44.44  (18.88-73.33) | 8.33  (0.43-35.39) | 60.00  (42.32-75.41) | 19.44  (9.75-35.03) |
| Cumulative to DPE 4 | 89.13  (76.96-95.27) | 75.00  (60.56-85.43) | 88.89  (71.94-96.15) | 81.48  (63.30-91.82) | 93.08  (89.32-95.58) | 77.99  (72.56-82.61) |
| Cumulative to DPE 6 | 98.15  (90.23-99.91) | 94.44  (84.89-98.09) | 96.55  (82.82-99.82) | 93.10  (78.04-98.09) | 95.38  (92.11-97.34) | 79.54  (74.21-84.00) |
| Cumulative to DPE 10 | 100.0  (93.36-100.0) | 96.43  (87.88-99.02) | 100.0  (88.97-100) | 93.55  (78.28-98.21) | 99.23  (97.24-99.79) | 82.24  (77.12-86.41) |
