## Supplemental Figures for "Performance of Rapid Antigen Tests Based on Symptom Onset and Close Contact Exposure: A secondary analysis from the Test Us At Home prospective cohort study"

### Slide 1
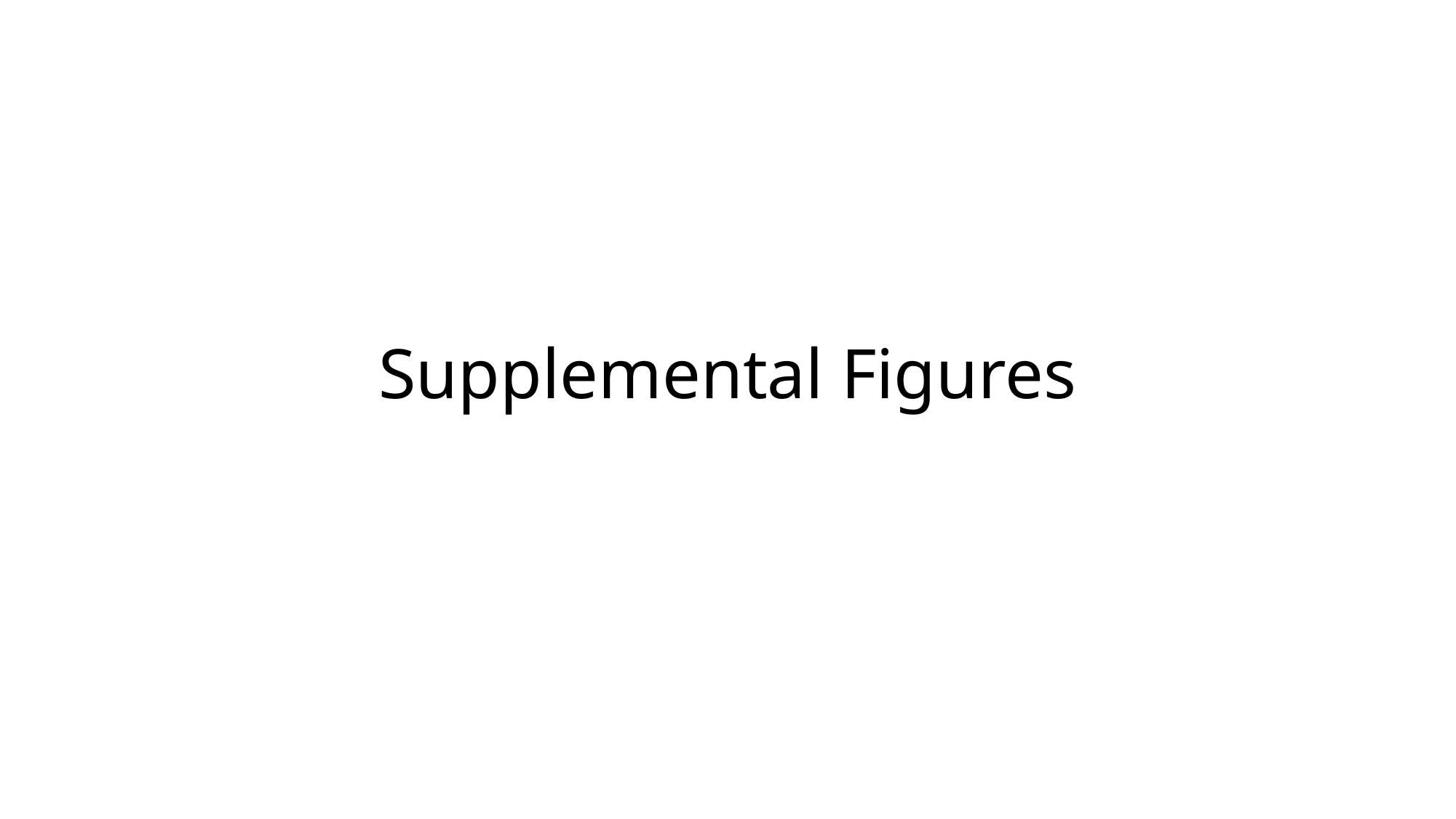

#### Supplemental Figures

### Slide 2
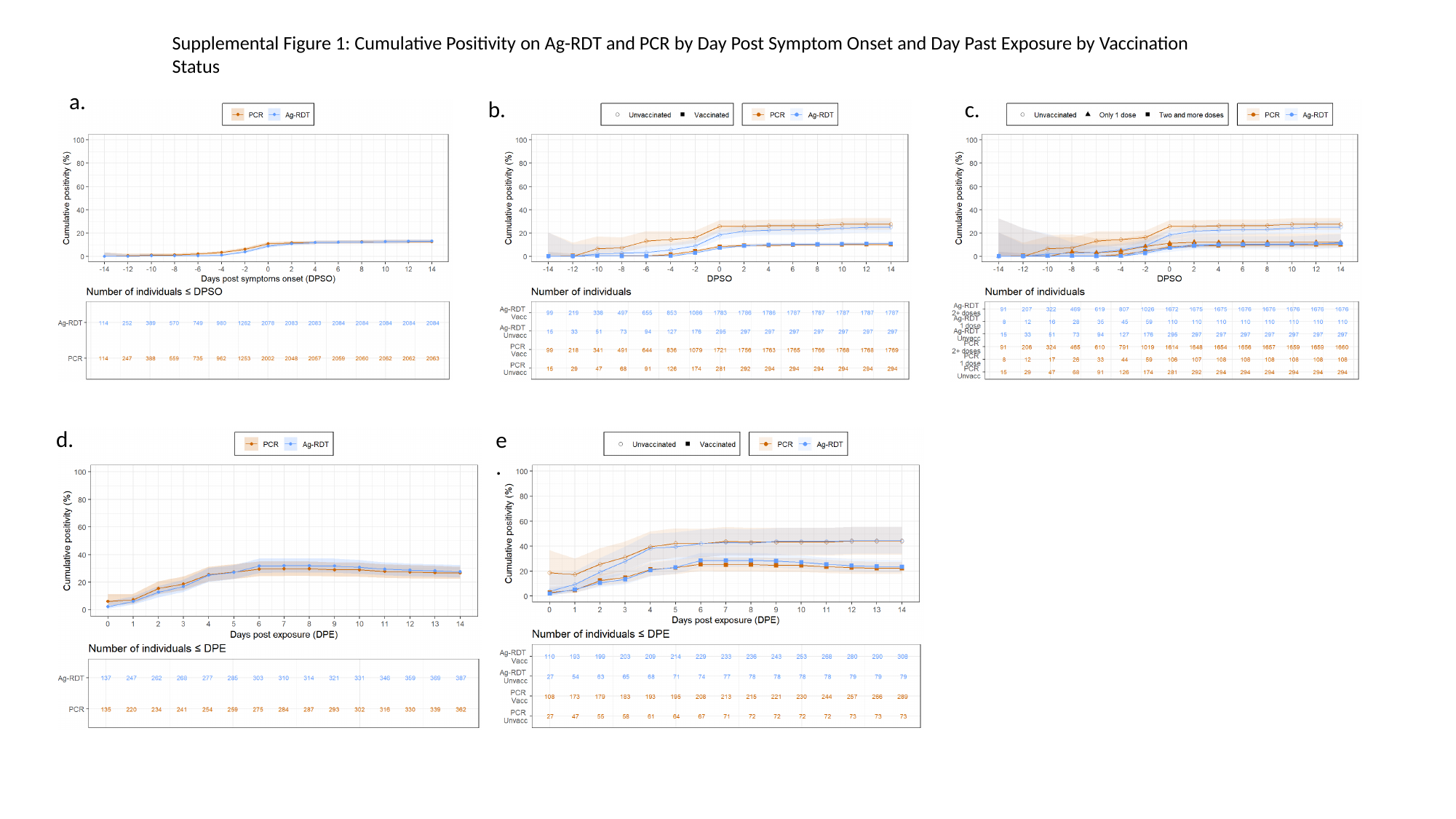

Supplemental Figure 1: Cumulative Positivity on Ag-RDT and PCR by Day Post Symptom Onset and Day Past Exposure by Vaccination Status
a.
b.
c.
d.
e.

### Slide 3
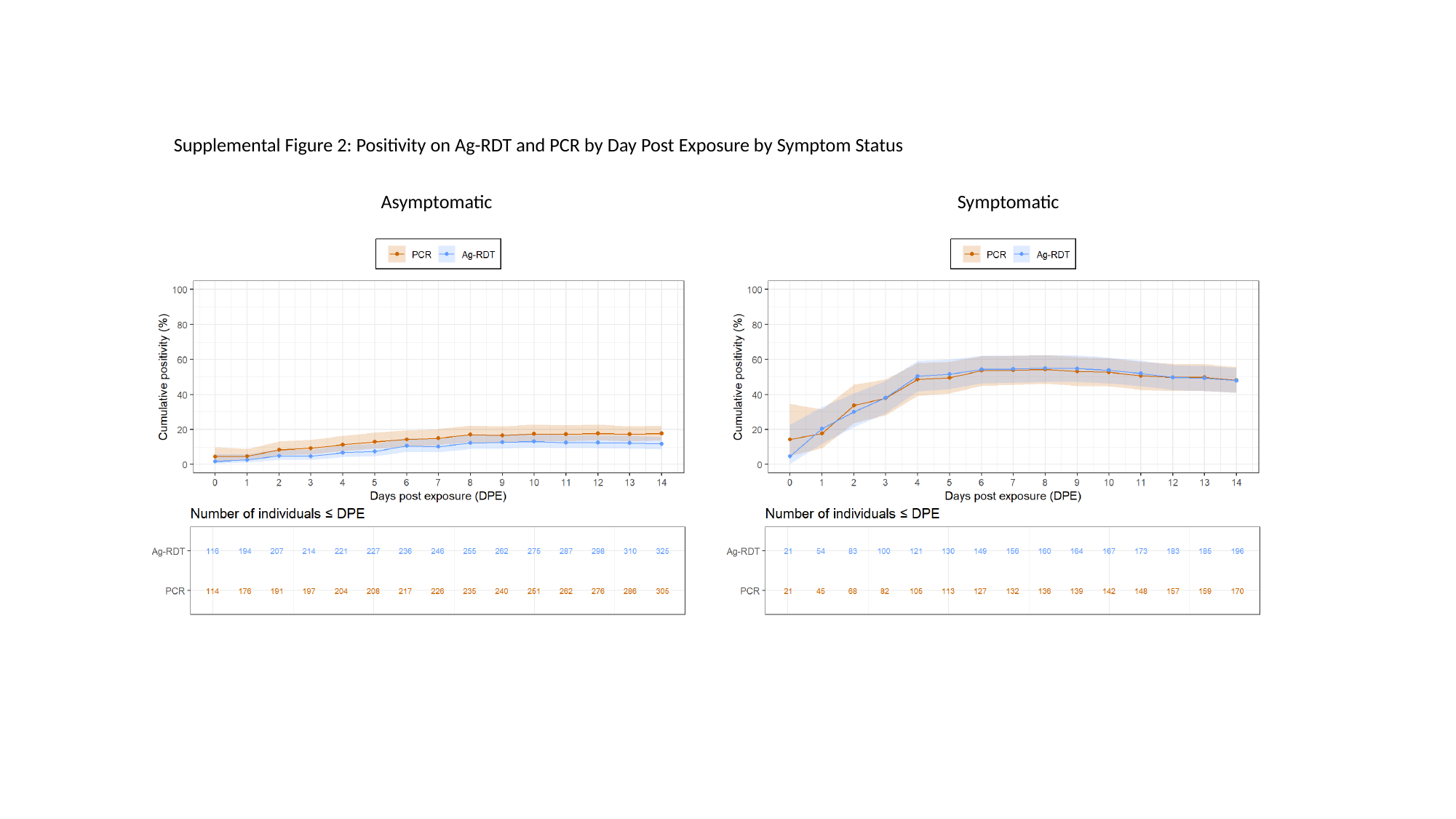

Supplemental Figure 2: Positivity on Ag-RDT and PCR by Day Post Exposure by Symptom Status
Asymptomatic
Symptomatic

### Slide 4
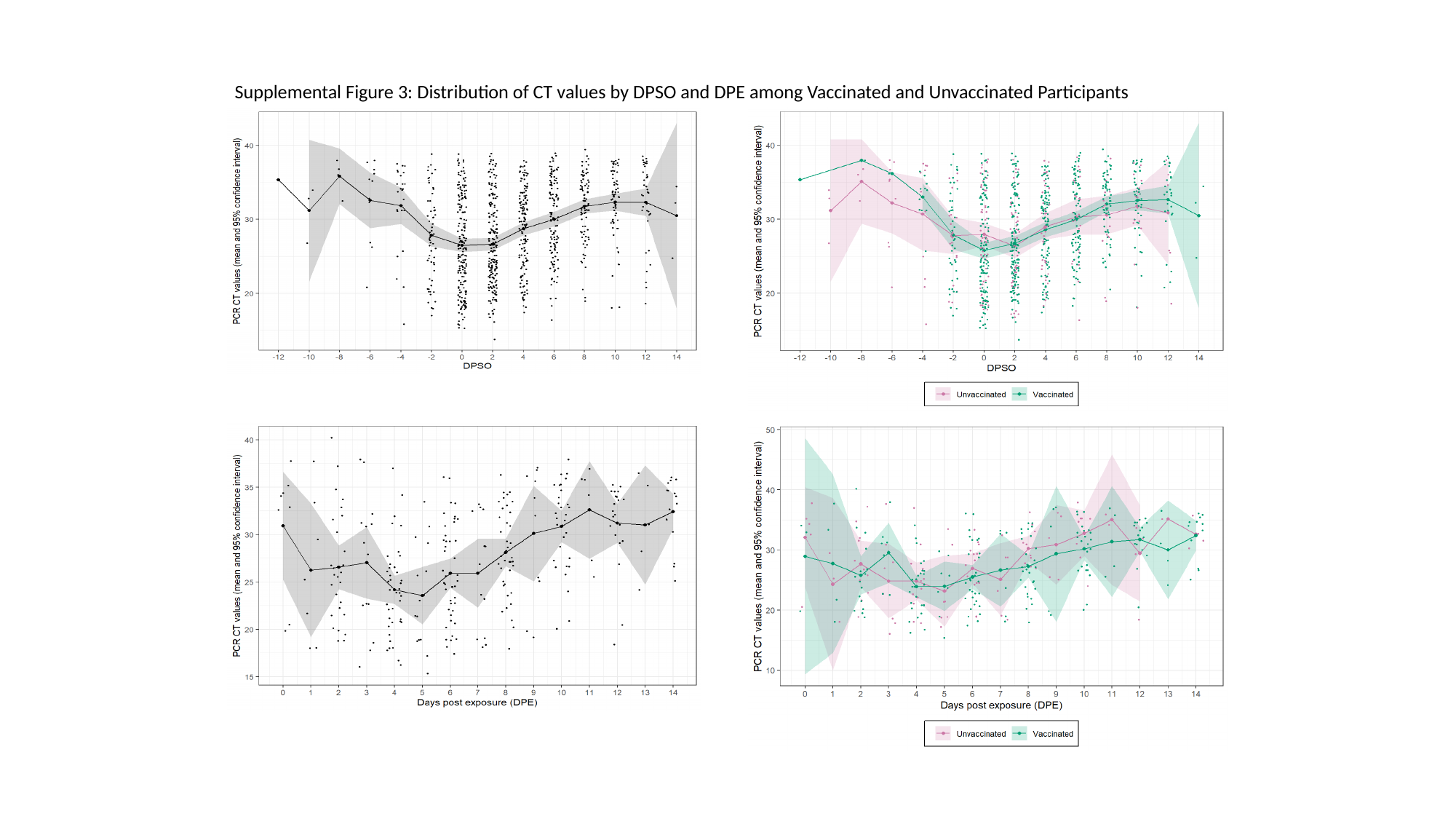

Supplemental Figure 3: Distribution of CT values by DPSO and DPE among Vaccinated and Unvaccinated Participants
